# Relationship of Body Composition with Cortisol and Dehydroepiandrosterone-Sulfate Levels in Korean Men and Women

**DOI:** 10.1101/2025.03.23.25324482

**Authors:** Sat Byul Park

**Affiliations:** Department of Family Practice and Community Health, Ajou University School of Medicine, Suwon, Korea

**Keywords:** Cortisol, DHEA-S, Cortisol/DHEA-S ratio, Body Composition, Central Obesity

## Abstract

**Objectives:** Glucocorticoids, mediated by the activation of the HPA axis, affect metabolic responses, insulin resistance, lipolysis and body fat distribution. Dehydroepiandrosterone-sulfate (DHEA-S) is a hormone produced by the adrenal glands and a precursor of sex hormones. The balance and interaction between cortisol and DHEA-S can significantly affect body composition. This study aimed to investigate the relationship between cortisol and DHEA-S levels, cortisol/DHEA-S ratio, and body composition in Korean men and women.

**Methods:** In total, 802 adults participated in this study between January 2018 and March 2023. Socio-demographic data and lifestyle factors were assessed using questionnaires. Body composition, clinical blood pressure, and metabolic variables, including cortisol and DHEA-S levels, were assessed. Cortisol and DHEA-S scores were analyzed in relation to height, body weight(BW), body mass index(BMI) and waist circumference(WC) according to age and sex.

**Results:** Participants had a mean age of 52.6±11.7 years. Cortisol levels adjusted for age and gender were negatively correlated with BW, WC and BMI. This result was more significant in women than in men. DHEA-S levels were positively correlated with height, BW and WC after adjusting for age. The cortisol/DHEA-S ratio was associated with lower height and BW after adjusting for gender. Logistic regression for cortisol, DHEA-S and the cortisol/DHEA-S ratio in the prediction of central obesity was significant for men after adjusting for age and BMI.

**Conclusions:** Elevated cortisol concentrations are associated with lower adiposity. DHEA-S levels were positively correlated with height and body mass. The prediction of central obesity was associated with cortisol and the cortisol/DHEA-S ratio in men and negatively associated with DHEA-S.

## INTRODUCTION

Glucocorticoids directly impair insulin sensitivity in adipocytes (1) and promote free fatty acid release from mature adipocytes through hormone sensitive lipase-mediated lipolysis (2). Abdominal obesity promotes insulin resistance and is considered a key part of metabolic syndrome (3, 4). Although cortisol has been implicated as a pathophysiological contributor to idiopathic obesity, circulating cortisol has not consistently been shown to be elevated in persons with obesity as compared to individuals with normal weight (5, 6). A recent study using both cross-sectional and longitudinal analysis to determine the associations between body composition and serum cortisol concentrations in men showed that weight change drives changes in cortisol concentrations, rather than vice versa (7).

Dehydroepiandrosterone-sulfate (DHEA-S) levels are abnormal in persons with insulin resistance syndrome (IRS): DHEA-S is decreased in men and increased in women. An association between low DHEA levels and increased cardiovascular mortality has been reported in men, whereas, in women, high DHEA-S levels are associated with a higher prevalence of cardiovascular disease (8). For men, hyperinsulinemia favors the reduction of adrenal androgen levels by inhibiting the enzyme 17-20 lyase, which determines androstenedione and DHEA levels without affecting glucocorticoid and mineralocorticoid production (9).

Among the various indices used to assess obesity, body mass index (BMI) is the most practical. However, BMI cannot distinguish between fat mass, lean mass, and adiposity types. Individuals with similar BMI may possess distinct body compositions, such that individuals with a high BMI may be physically fit with a low mortality risk compared to individuals with low BMI but malfunctioning adiposity (10, 11). The type of adiposity seems to play a role in the obesity paradox. Indices such as waist circumference (WC), waist-to-hip ratio, and body fat percentage (%BF) should better reflect body fatness (12).

The secretion of cortisol and DHEA-S in an appropriate balance is essential for the maintenance of biological function and homeostasis. A high cortisol/DHEA-S ratio is reportedly associated with mortality (13), dementia (14), metabolic syndrome (15), and reduced immunity following physical stress (16). Furthermore, discordant secretion of these two adrenal hormones is related to aging (17, 18). Mechanisms linking hormones such as cortisol and DHEA-S with IRS and body composition are not completely understood; additionally, there are still many controversial aspects. This study aimed to assess the relationship between cortisol, DHEA-S levels, and the cortisol/DHEA-S ratio with body composition in healthy individuals.

## Materials and Methods

### Study Population

A total of 1,341 Korean adults aged >20 years who underwent medical evaluations at Ajou University Hospital between January 2018 and March 2023 were enrolled in this study. Participants were excluded if they were taking steroid medication or did not provide information about their body composition (N = 539). The final sample size was 802 participants (71.4 % women).

### Institutional review board statement

The study protocol was inspected and approved by an independent Institutional Review Board in Ajou University Hospital, which reviewed the ethical issues pertaining to studies on human participants (AJOUIRB-DB-2025-102). In addition, this study was a retrospective study of medical records and all data were fully anonymized before I accessed them. The requirement for informed consent was waived by the IRB committee.

### Data Collection

Sociodemographic characteristics, smoking status, alcohol consumption, and activity level were assessed using questionnaires. Height and weight were measured using bioelectrical impedance analysis (Inbody 3.0, Biospace, Korea, 2021) after overnight fasting. BMI was calculated as weight divided by height squared (kg/m^2^) (19). WC was measured between the lower rib and iliac crest by a trained nurse. Blood pressure was measured using a semiautomated blood pressure monitor (TM-2650A; PMS Instruments, Tokyo, Japan) after resting for at least 15 min. Venous blood samples were collected after 12-h overnight fasting and 24 h of abstinence from vigorous physical activity. Cortisol and DHEA-S levels were assessed using Radio Immuno Assay (RIA) (Diagnostic Product Cooperation, CA, USA).

### Statistical Analyses

Nominal variables were analyzed using chi-square analysis, and Student’s t-test was used to compare the mean values of general characteristics and laboratory test results between men and women. Partial correlations were calculated to adjust for age and sex. Logistic regression analysis was used to evaluate the serum levels of cortisol and DHEA-S, and the cortisol/DHEA-S ratio for the prediction of central obesity. Statistical significance was set at P<0.05. SPSS (version 29.0) was used for all statistical analyses.

## Results

### Demographic characteristics

The study population consisted of 802 individuals, including 229 men (29.6 %) and 573 women (71.4 %) (Table 1). The mean age and BMI were 52.6±11.7 years and 24.2±4.1 kg/m^2^, respectively. The mean cortisol and DHEA-S levels were 12.6±4.5 μg/dL and 104.5±80.7 μg/dL, respectively. Mean levels (± Std) of study variables are presented by sex in Table 1. Sex differences were observed in several demographic and clinical variables, including WC, BMI, cortisol and DHEA-S levels and the cortisol/DHEA-S ratio.

### Correlations between cortisol, DHEA-S, cortisol-DHEA-S ratio, and body composition

The partial correlation analyses evaluating the association between cortisol levels and body composition are shown in Table 2. Cortisol levels adjusted for age and sex were negatively correlated with body weight (BW), WC, and BMI (P=0.004, P=0.023, and P=0.003, respectively). This result was more significant in women than in men.

The associations between DHEA-S level and height, BW, and WC were significant (P=0.000, P=0.000, and P=0.018, respectively) after adjusting for age (Table 3).

The cortisol/DHEA-S ratio was negatively associated with height after adjusting for sex (P=0.000). The cortisol/DHEA-S ratio was also associated with a low BW (P=0.043) (Table 4). No association was observed between the cortisol/DHEA-S ratio and other body compositions.

### Logistic regression analysis for identification of factors predicting central obesity

In men, higher cortisol levels were associated with an increased risk of central obesity after adjusting for age and BMI (P=0.009) (Table 5). In contrast, DHEA-S level was negatively associated with an increased risk of central obesity (P=0.011) (Table 6). The cortisol/DHEAS ratio was associated with central obesity (P=0.02) (Table 7). However, this effect was not statistically significant among women.

## Discussion

This study showed that cortisol levels and cortisol/DHEA-S ratio were negatively correlated with body composition. In contrast, DHEA-S levels were positively correlated with body composition. In men, higher cortisol levels and cortisol/DHEA-S ratios were associated with an increased risk of central obesity. DHEA-S levels were negatively associated with an increased risk of central obesity. Cortisol itself was associated with neuroendocrine abnormalities such as inhibited gonadal function, low growth hormone levels, and increased sympathetic nervous system activity. However, these findings have not been confirmed in women (20), although a correlation between low plasma cortisol levels and central obesity has been reported (21).

The association between low cortisol levels and obesity has been explained by a hypothalamus-pituitary-adrenal gland axis disorder that causes an inversion of the normal circadian variation of cortisol with low morning and high afternoon levels. In addition, a high renal cortisol clearance rate has been reported, suggesting that it may have been developed to compensate for increased cortisol secretion and maintain low plasma cortisol levels to avoid its deleterious effects on target organs. There is considerable uncertainty concerning the significance of this association in women (21). Results of the present study suggest that cortisol levels and cortisol/DHEA-S ratio are negatively correlated with body composition.

There are still contradictory data regarding the relationship between DHEA-S and body fat accumulation, and while some studies have reported no association (22) or others did find an inverse relation between the hormone and obesity, such as anti-obesity effect of DHEA-S through the *in vitro* stimulation of adipose tissue lipolysis (23, 24). Differences in the age of study participants may have represented an important confounding factor in the study of the relationships between obesity, body fat distribution, and endogenous DHEA-S (25). One study found a statistically significant association between DHEAS and percent body fat, total adipose mass, and percent lean body mass in men (26).

The mechanisms linking obesity, body fat distribution, and serum DHEA or DHEAS levels have not been fully elucidated, although several possibilities have been proposed. Gordon et al. observed that cell conversion to mature adipocytes is impeded in the presence of DHEA, whereas DHEA-S exhibits a minor inhibitory effect on glucose-6-phosphate dehydrogenase activity (27). The role of DHEA as a precursor for active androgens and estrogens may also play a role in the relationship between obesity, body fat distribution, and circulating DHEA. Steroids are lipophilic compounds that are highly soluble in adipose tissue (28, 29). Moreover, adipose tissue expresses several steroid-converting enzymes necessary for the local synthesis of active androgens/estrogens from inactive precursors such as DHEA (28, 30, 31). The enzymes responsible for the inactivation of androgens/estrogens are also present in adipose tissue (32), thereby modulating the intracellular levels of active steroids. The intracrine conversion of DHEA to androgens or estrogens in a site-specific manner could affect adipocyte physiology and modulate adipose tissue accumulation and mobilization (31).

Although DHEA and DHEA-S levels decline in both men and women with age (33), testosterone levels are generally well maintained in men into old age and are always several-fold higher than the levels observed in women. There was a negative association between cortisol level and BMI and WC in women, and no association between cortisol level and BMI or WC in men. These findings are consistent with previous studies showing that cortisol levels are not elevated in individuals with obesity as compared to individuals with normal weight (5, 6), and are also in agreement with a study suggesting diminished stimulability of the hypo-thalamo-pituitary (HPA) axis in individuals with obesity (34). Cortisol is released from subcutaneous adipose tissue by 11beta-HSD1 in humans, and increased enzyme expression in obesity is likely to increase local glucocorticoid signaling and contribute to whole-body cortisol regeneration (35). However, because the previous findings are not entirely consistent, whether enzymatic overactivity is a cause or a result of obesity remains unclear.

The study population of this study comprised patients who visited a University Hospital, which may have limited the generalizability of our findings. Second, because this study employed a cross-sectional design, it was not possible to determine whether cortisol and DHEAS levels were causally related to changes in body composition or the development of central obesity. Longitudinal studies could assess whether circulating cortisol and/or DHEA-S concentrations increase changes in body composition and the risk of developing central obesity. In addition, variables that could influence body composition, especially in women, such as hormone therapy, were not considered. Further research on the mechanisms underlying these observations may lead to new therapeutic approaches to reduce changes in body composition and central obesity.

## Conclusion

This study demonstrates that plasma cortisol and DHEA-S concentrations contribute to the effect of body composition in Korean adults and suggests a relationship between HPA activity, as indicated by increased fasting DHEA-S concentration, and increased central obesity, independent of adiposity in Korean men.

## Data Availability

The data that support the study are available upon reasonable request to the corresponding author.

